# Modelling the Potential Health Impact of the COVID-19 Pandemic on a Hypothetical European Country

**DOI:** 10.1101/2020.03.20.20039776

**Authors:** Nick Wilson, Lucy Telfar Barnard, Amanda Kvalsvig, Ayesha Verrall, Michael Baker, Markus Schwehm

## Abstract

A SEIR simulation model for the COVID-19 pandemic was developed (http://covidsim.eu) and applied to a hypothetical European country of 10 million population. Our results show which interventions potentially push the epidemic peak into the subsequent year (when vaccinations may be available) or which fail. Different levels of control (via contact reduction) resulted in 22% to 63% of the population sick, 0.2% to 0.6% hospitalised, and 0.07% to 0.28% dead (n=6,450 to 28,228).

---

There is pandemic spread of the new coronavirus “SARS-Cov-2”, causing the disease “COVID-19”, with the World Health Organization (WHO) reporting over 200,000 cases and over 8000 deaths on 19 March 2020 [1]. One approach to informing the potential health burden and relevant control measures for a new pandemic is to study its dynamics using mathematical models. Recently published mathematical modelling work on COVID-19 has reported that “in most scenarios, highly effective contact tracing and case isolation is enough to control a new outbreak of COVID-19 within 3 months” [2]. Another modelling study found that “combining all four interventions (social distancing of the entire population, case isolation, household quarantine and school and university closure) is predicted to have the largest impact, short of a complete lockdown which additionally prevents people going to work” [3]. Other such models have been used to estimate the impact of disease control measures in China [4, 5]. Given this background, we explore the potential health impact of the spread of the COVID-19 pandemic in a hypothetical European country of 10 million people to determine the potential impact of control measures, particularly to push the epidemic into a subsequent year, a time when a vaccine might become available.

Europe’s first cases of COVID-19 were reported to WHO on 25 January in France [6]. But 19 March there were 52 of these countries with reported cases, with 12 reporting 1000+ cases [1]. By this time major control interventions were in place in many of these countries [7].

In this modelling, we took the standard approach of using a deterministic SEIR model i.e., key compartments for: susceptible [S], exposed [E], infected [I], and recovered/removed [R]. We developed this model specifically for COVID-19. It is freely available online with a dashboard display to facilitate user interaction (http://covidsim.eu; version 1.0, 19 March). The Appendix details the parameters, derived variables and differential equations used in the CovidSIM model. Table A1 in the Appendix provides the input parameters used in the model, as based on available publications and best estimates used in the modelling work on COVID-19 to date (as known to us on 18 March 2020).

Our results suggest that pushing the peak of the epidemic into the next year, when assuming a low basic reproduction number (R_0_) of only 1.5, was achievable with “general contact reduction” at levels of over 11% to over 27% (for differing time periods) (Table 1). Similarly, it was achievable for probabilities of isolating symptomatic cases in hospital in the range of over 18% to over 41% (Table 1). Using instead a possibly more realistic value of 2.5 for R_0_, the only way of shifting the epidemic peak into the next year with interventions lasting for nine months, was if: (i) over 65% of contacts were reduced; or (ii) if the probability of cases being isolated in hospitals was extremely high at over 98%. Also for R_0_ = 3.5, pushing the epidemic peak into the subsequent year was generally not possible, except if contact reduction was over 61% for the rest of the simulated year.

**Table 1:** Threshold analyses for pushing the peak of the COVID-19 epidemic in the hypothetical European country into the next year (i.e., pushing the peak to after day 365 of the simulation with the start of the simulation on 15 February 2020, the date we assumed that the infection spread began)

| Intervention settings | Assumed basic reproduction number |  |  |
| --- | --- | --- | --- |
| | $R_0=1.5$ | $R_0=2.5$ | $R_0=3.5$ |
| ***Intensity and length of “general contact reduction” starting on day 5 of the simulation*** |  |  |  |
| Level of general contact reduction for 6 months needed to push the epidemic into the next year | >27% | Not possible | Not possible |
| – for 9 month intervention period (274 days) | >14% | >65% | Not possible |
| – for rest of the simulated year | >11% | >46% | >61% |
| ***Proportion of cases in hospital isolation (with home isolation at 50% effectiveness when hospital capacity is exceeded; beginning on day 5 of the simulation)*** |  |  |  |
| Probability of case isolation in hospital needed to push the epidemic into the next year (for a 6 month intervention period) | >41% | Not possible | Not possible |
| – for 9 month intervention period (274 days) | >21% | >98% | Not possible |
| – for rest of the simulated year | >18% | >78% | Not possible |

The feasibility of achieving high levels of contact reduction and case isolation is very uncertain – especially around sustaining these for long periods of time. At least in the short-term China has used intensive containment measures successfully as per the findings of the WHO-China Joint Mission Report [8]. This report stated that: “China has rolled out perhaps the most ambitious, agile, and aggressive disease containment effort in history.” While it is an open question around the generalisability of the Chinese approach to other jurisdictions [9], there is also evidence of containment success (as of late March 2020) outside mainland China, from Singapore, Hong Kong and Taiwan [10]. But for European countries currently with thousands of cases, it may be too late to adopt such intensive containment approaches and the best approach may simply be to apply less intense control measures that balance minimising health loss with minimising social and economic disruption (while also putting major resources into rapid vaccine development).

Figure 1 shows the scenarios at three different levels of R_0_ when combined with “25% general contact reduction”. At R_0_ = 3.5 the epidemic still peaks during the intervention period whereas for R_0_ = 2.5 the peak is pushed into the post-intervention period. For R_0_ = 1.5 the peak is nearly pushed into the subsequent year.

**Figure 1:**
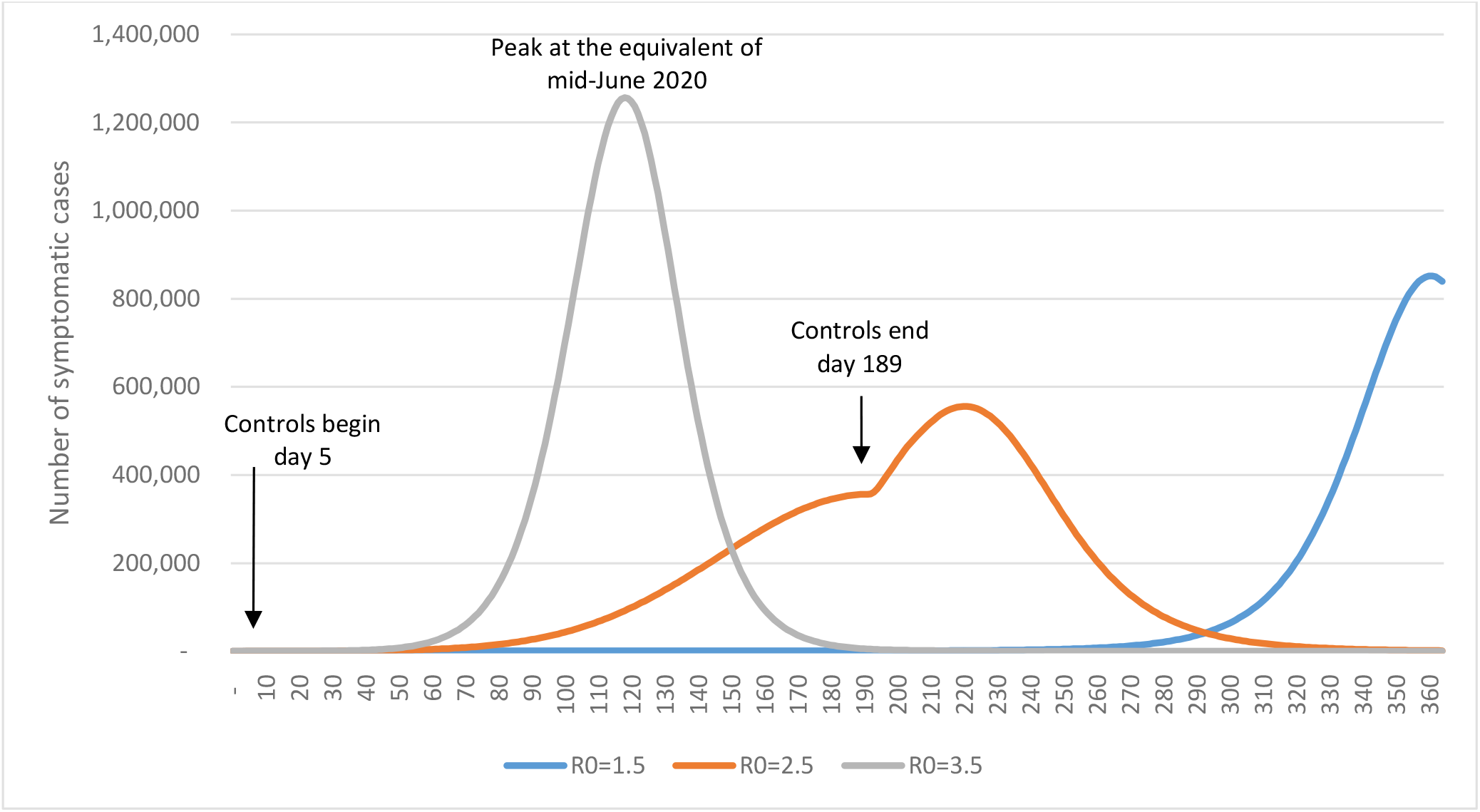
Numbers of prevalent symptomatic COVID-19 cases by day for the R_0_ = 1.5, 2.5, and 3.5 scenarios and all with 25% of “general contact reduction” as modelled using CovidSIM (unrecognised introduction of the infection occurs at time 0; control measures begin at day 5 and end six months later i.e., on day 189).

Figure 2 again demonstrates the importance of the timing of the intervention period. A higher level of contact reduction (at 50% vs 25%) more effectively suppresses the epidemic in the intervention period – but when the intervention period ends it causes a higher epidemic peak and a higher total health burden than for the 25% reduction (Table 2).

**Table 2:** Estimated health impacts from COVID-19 in a hypothetical European country of population 10 million using CovidSIM for a range of basic reproduction number (R_0_) values and differing intensity of “general contact reduction” as the control measure (see Table A1 for input parameters)

| Key results | $R_0=1.5^*$ | | $R_0=2.5$ | | $R_0=3.5$ | |
| --- | --- | --- | --- | --- | --- | --- |
|  | 25% control for 6 months | 50% control for 6 months | 25% control for 6 months | 50% control for 6 months | 25% control for 6 months | 50% control for 6 months |
| General pattern seen for symptomatic cases | Peak late in the year | Peak in next year | 1 peak after intervention period | 1 peak after intervention period | 1 peak in intervention period | 1 peak after intervention period |
| **Symptomatic cases (which are 67% of all infected cases)** |  |  |  |  |  |  |
| Total | 3,327,382 | 2,200,102 | 5,415,805 | 6,225,554 | 5,521,618 | 6,272,963 |
| Proportion of population (%)** | 33.3% | 22.0% | 54.2% | 62.3% | 55.2% | 62.7% |
| Peak week for incidence | 52 | Next year | 32 | 40 | 17 | 32 |
| Peak month for incidence | 12 | Next year | 7 | 9 | 4 | 7 |
| Number of sick people on the worst day of the simulated year | 851,592 | 766,739 | 555,676 | 1,743,927 | 1,256,488 | 1,494,048 |
| Proportion of population sick on the worst day (%)** | 8.5% | 7.7% | 5.6% | 17.4% | 12.6% | 14.9% |
| **Consultations (40% of symptomatic cases seek consultations, possibly mainly telephone/internet)** |  |  |  |  |  |  |
| Total | 1,330,953 | 880,041 | 2,166,322 | 2,490,222 | 2,208,647 | 2,509,185 |
| Proportion of population (%)** | 13.3% | 8.8% | 21.7% | 24.9% | 22.1% | 25.1% |
| **Severe cases likely to require hospitalisation (1.0% of symptomatic cases)** |  |  |  |  |  |  |
| Total | 33,274 | 22,001 | 54,158 | 62,256 | 55,216 | 62,730 |
| Proportion of population (%)** | 0.3% | 0.2% | 0.5% | 0.6% | 0.6% | 0.6% |
| Number of people in hospital on the worst day (if capacity existed) | 8,516 | 7,667 | 5,557 | 17,439 | 12,565 | 14,940 |
| Proportion of population in hospital on the worst day (%)** | 0.09% | 0.08% | 0.06% | 0.17% | 0.13% | 0.15% |
| **Cases likely to require ICU (25% of hospitalised cases)** |  |  |  |  |  |  |
| Total | 8,318 | 5,500 | 13,540 | 15,564 | 13,804 | 15,682 |
| People in ICU on the peak day (if capacities exist) | 2,129 | 1,917 | 1,389 | 4,360 | 3,141 | 3,735 |
| **Cases likely to require ventilation in ICU (50% of those in ICU)** |  |  |  |  |  |  |
| Total** | 4,159 | 2,750 | 6,770 | 7,782 | 6,902 | 7,841 |
| **Deaths (case fatality risk amongst symptomatic cases of 0.45%)** |  |  |  |  |  |  |
| Total | 11,194 | 6,450 | 24,365 | 28,014 | 24,847 | 28,228 |
| Proportion of population (%)** | 0.11% | 0.07% | 0.24% | 0.28% | 0.25% | 0.28% |

\* Results for $R_0=1.5$ scenarios are shaded as they were both right truncated (i.e., only the results for the first 365 days of the simulation are reported) as the epidemic peak was pushed into either very late in the year (for 25% reduction) or the following year (for 50% reduction).
\*\* Results in these rows were not standard outputs for the CovidSIM model but were based on further Excel-based calculations from the CovidSIM output.

**Figure 2:**
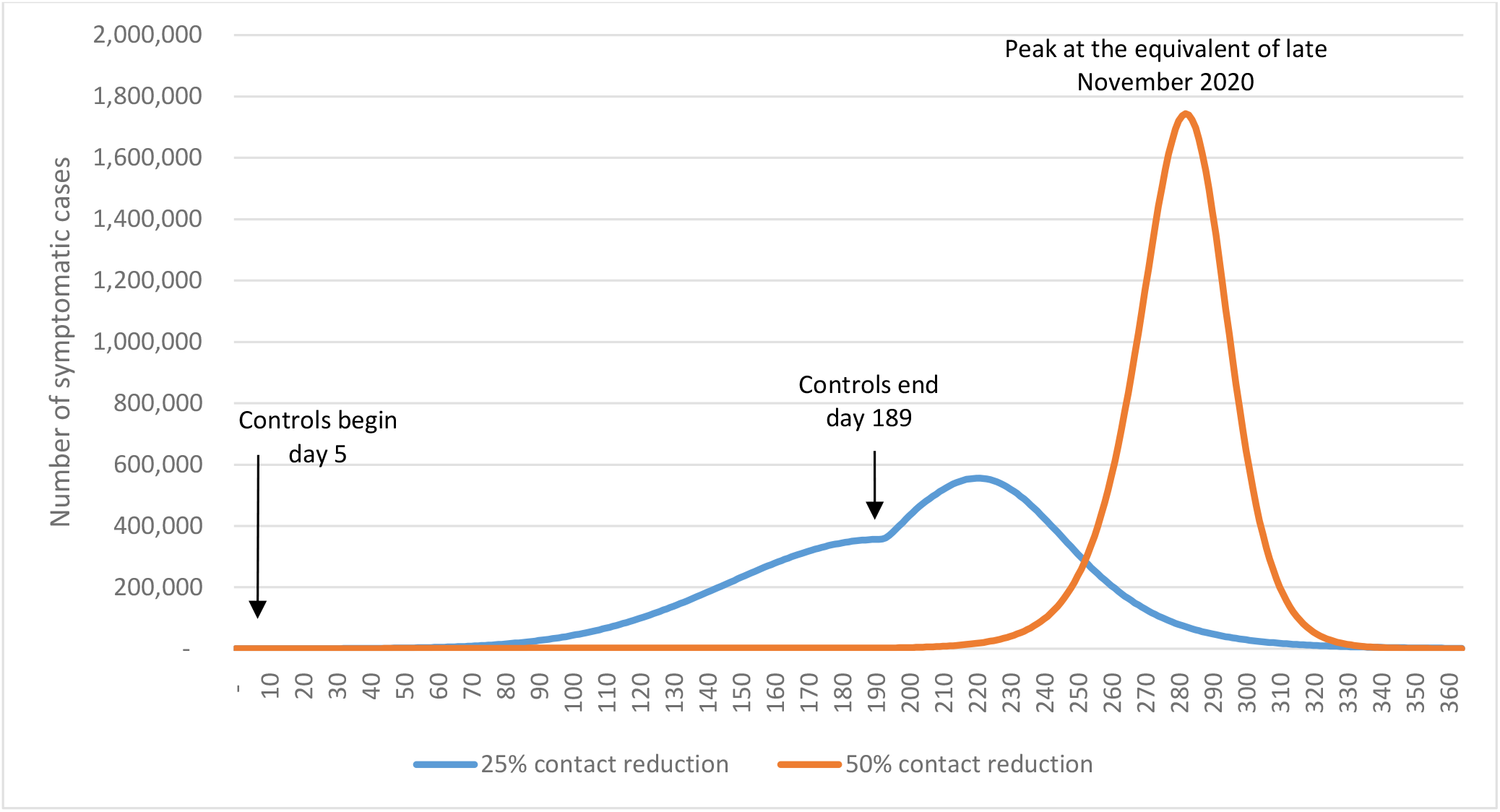
Numbers of prevalent symptomatic COVID-19 cases by day for the R_0_ = 2.5 scenarios and different levels of general contact reduction as modelled using CovidSIM (unrecognised introduction of the infection occurs at time 0; control measures begin at day 5 and end six months later i.e., on day 189).

Considering the three levels of R_0_ and the two levels of “general contact reduction” (at 25% and 50%) resulted in: 22% to 63% of the population sick, 8.8% to 25.1% seeking a medical consultation, 0.2% to 0.6% needing to be hospitalised, 5,500 to 15,682 people needing critical care (in an ICU), 2,750 to 7,841 requiring ventilators, and 6,450 to 28,200 dying (0.07% to 0.28% of the population) (Table 2). The worst scenario with 0.28% of the population dying, compares to Europe in the 1918 influenza pandemic where 1.1% died (2.64 million excess deaths) [11].

Based on the recent age distribution data from China [12], these hospitalisations and deaths from COVID-19 would particularly occur amongst older age-groups. Overall, the levels of health service demand in the more severe scenarios would be completely unprecedented for a modern European country.

This is one of the first SEIR modelling studies of this new pandemic disease COVID-19 and the associated online simulation tool has advanced dashboard features and graphic visualisation of results that facilitate user engagement. Nevertheless, there are always limitations with modelling work. Some of these are discussed under issues with parameter uncertainty in Table A1, but here we note some of the more substantive limitations:

- There is still a high degree of uncertainty around many aspects of COVID-19 epidemiology. For example, the value of R_0_ could conceivably be higher than the highest level in the scenarios we modelled (at R_0_ = 3.5). Also, the case fatality risk (CFR) could be underestimated, especially if there was health system overload.
- The model was deterministic and not stochastic, though we largely offset this issue with modelling a wide range of scenarios. The lack of stochastic considerations mainly translates into increased uncertainty in the very early stages of epidemic spread, which then impacts on the timing of the peak.
- The model neither considers any long-term health damage to survivors (especially among risk patients) nor does it consider the hard-to-estimate health loss arising from untreated other health conditions as a result of having an overburdened health system. Likewise we do not consider the additional health harm to the health workers involved e.g., adverse mental health impacts arising from working during a pandemic [13, 14].

Some of these issues can be addressed when improved data becomes available on the epidemiology of COVID-19. Nevertheless, public health workers can consider using this model in their own countries and adapt the parameters according to local settings and as the epidemiological characteristics of COVID-19 are better ascertained.

## Data Availability

All the necessary data can be generated by the users themselves since the model is freely available online with a data file export capacity.

## Appendix: Description of the CovidSIM model and model input parameters

### Model dynamics

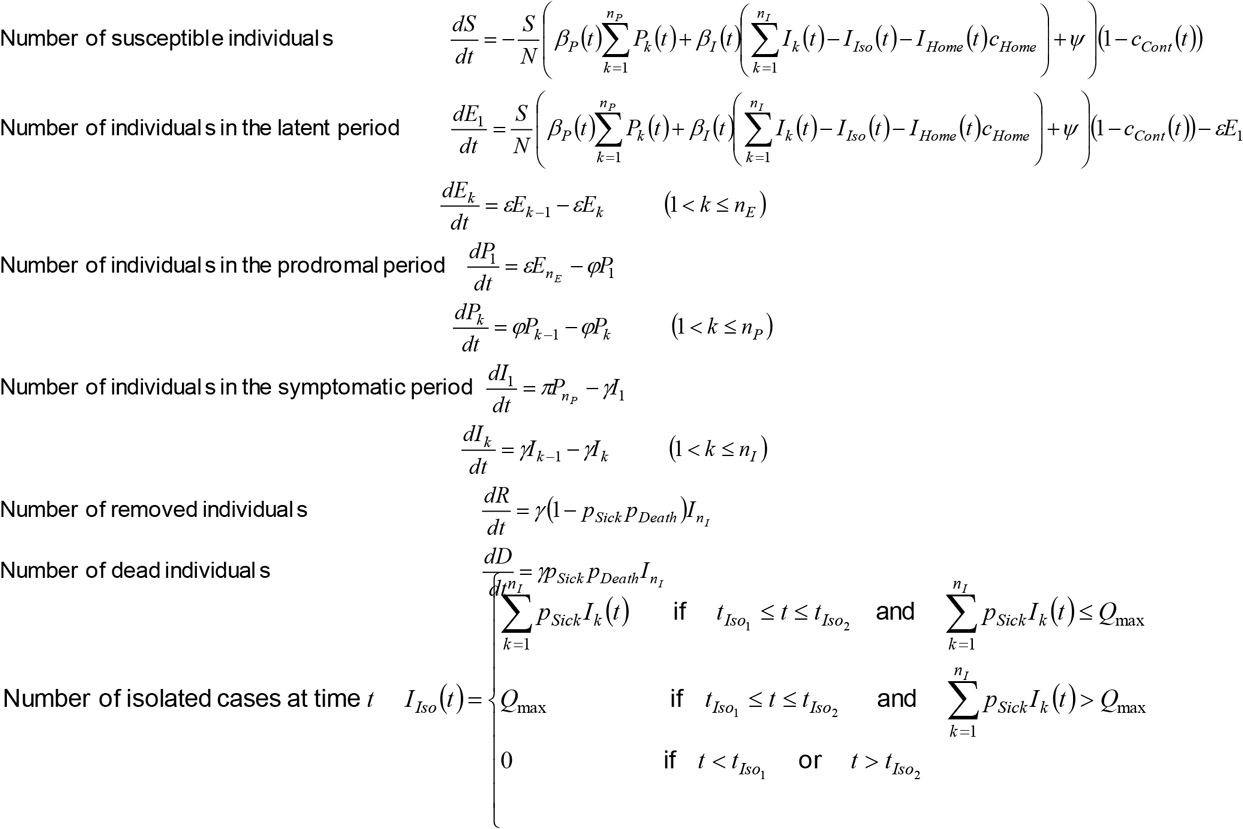

Number of fully isolated cases at time *t*

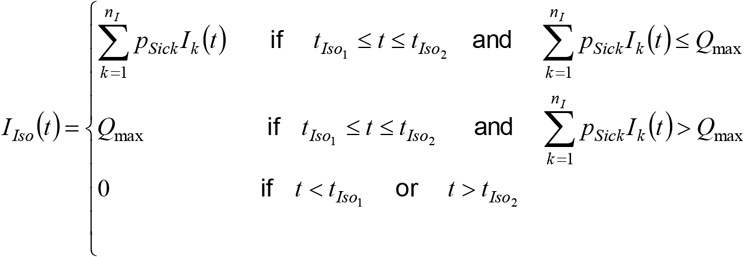

Number of home isolated cases at time *t*

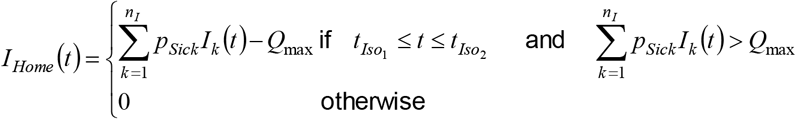

### Initial values

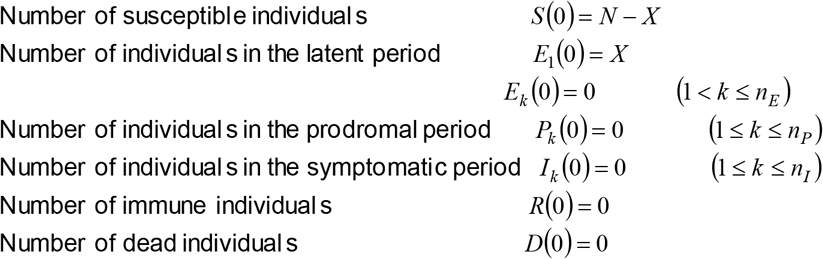

#### Parameters

*N*: Population size
*X*: Number of initial infections
*t*_max_: Day after introducti on of the infection when the transmissi on potiential is highest
*Q*_max_: Maximum isolation capacity
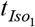: Time at which isolation measures start
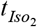: Time at which isolation measures end
*c*_*Home*_: Fraction of contacts which are prevented for cases who are in home isolation
*c*_*Cont*_: Fraction of contacts which are prevented
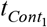: Time at which contact reduction starts
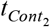: Time at which contact reduction ends
*c*(*t*): Fraction of contacts which are reduced at time *t*
*y*: Force of infection which originates fromoutside of the population (e.g. via travellers)
*R*_0_: Average value of the basic reproduction number
*a*: Amplitude of the seasonal fluctuatio n of *R*_0_
*D*_*E*_: Average duration of the latent period
*n*_*E*_: Number of stages for the latent period
*ε*: Stage transition rate in the latent period (*ε* = *n*_*E*_ / *D*_*E*_)
*D*_*P*_: Average duration of the prodoromal period
*n*_*P*_: Number of stages for the prodromal period
*φ*: Stage transition rate in the prodromal period (*φ* = *n*_*P*_ / *D*_*P*_)
*i*_*P*_: Relative infectiousness during prodromal period
*D*_*I*_: Average duration of the symptomatic period
*n*_*I*_: Number of stages for the symptomatic period
*γ*: Stage transition rate in the symptomatic period (*γ* = *n*_*I*_ / *D*_*I*_)
*β*_*I*_ (*t*): Effective contact rate of individual s in the symptomatic period at time *t β*_*I*_ (*t*) = *R*_0_/(*i*_*P*_*D*_*P*_ + *D*_*I*_)× (1+ *a* cos(*t* / 365))
*β*_*P*_ (*t*): Effective contact rate of individual s in the prodromal period at time *t* (*β*_*P*_ (*t*) = *β*_*I*_ (*t*)*i*_*P*_)
*p*_*Sick*_: Fraction of infected individual s who become sick
*p*_*Consult*_: Fraction of sick individual s who seek medical help
*p*_*Hosp*_: Fraction of sick individual s who are hospitalized
*p*_*ICU*_: Fraction of hospitalized individual s who are admitted to the ICU
*p*_*Death*_: Fraction of sick individual s who die from the disease

### Derived variables

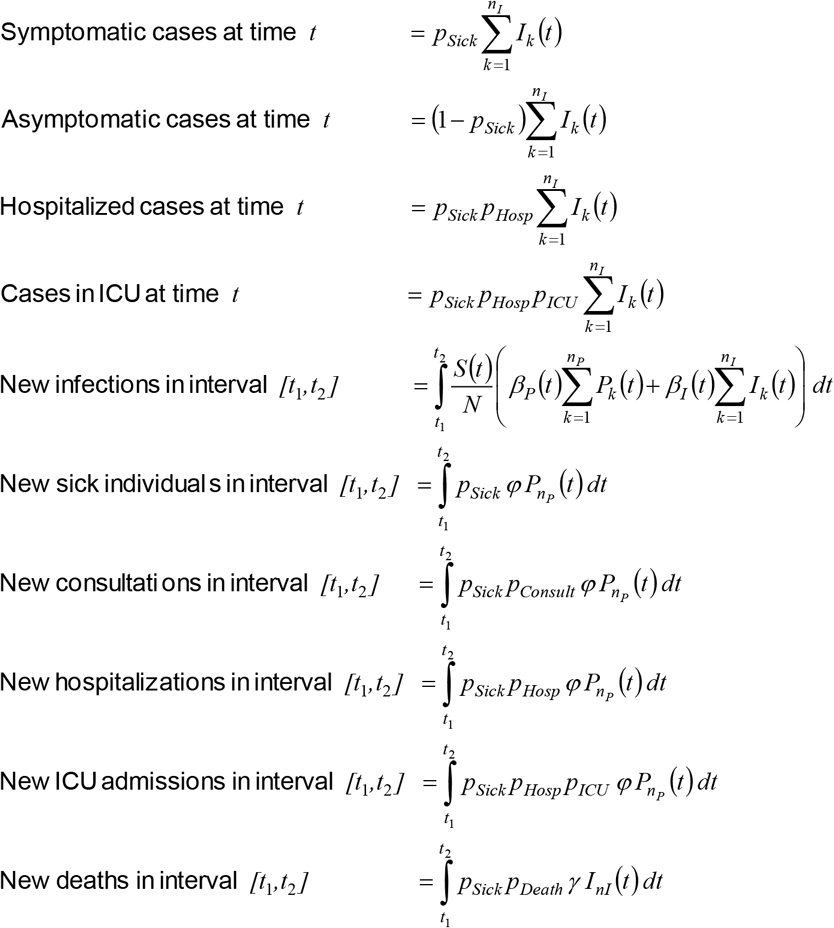

### Detection probability

SARS-CoV-2 infections which are brought into the country may not be detected and may spread without being noticed because the symptoms of COVID-19 may easily be confused with other influenza-like illnesses (ILI). Few practitioners may decide to order a SARS-CoV-2 test for what they regard a normal ILI patient while no community-transmitted cases in the population have been reported. If we assume that fraction of *p*_*Test*_ ILI patients who (a) seek medical help or who (b) are hospitalized or who (c) die from the disease are tested for SARS-Cov-2, then the probability that *not one single test* has been performed on a COVID-19 patient by time *t* despite the ongoing transmission in the population is given by:

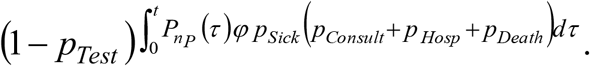

The probability that at least one test has been performed (and has returned a positive result) is then

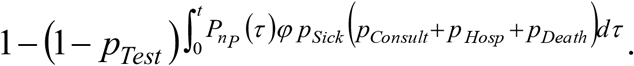

## Parameters used in the modelling with CovidSIM

**Table A1:** Input parameters for modelling the health impacts for a hypothetical European country using CovidSIM

| Parameter | Value/s used | Further details |
| --- | --- | --- |
| Population size | 10 million | The population we selected for a hypothetical European country. |
| Incoming infected people from outside of the study population | 10 per day (from 15 February) | We assumed that the first cases with subsequent silent transmission began on 15 February when nine European countries had cases. For the whole course of the one year simulation, we assumed an average of 10 infected people per day were arriving in the hypothetical country from other countries (e.g., by legal and/or illegal crossing of land borders). |
| Infections that lead to sickness | 67% | This figure is still uncertain but we used the same estimate as per modelling by Imperial College at “two thirds of cases being sufficiently symptomatic to self-isolate” [3]. Of note is that another modelling study used a 50% value [15]. Nevertheless, some proportion of asymptomatic cases is consistent with the findings of a very large Chinese study [12], where 81% of cases of COVID-19 did not involve severe illness. |
| Sick people seek medical help (including telephone and internet consultations) | 40% | We used the default value in the CovidSIM model, which is based on medical consultations for influenza-like illness (ILI). During a pandemic there might be a shift away from face-to-face consultations with health workers, so that some of these consultations may be either telephone or internet-based. This parameter is not used for determining subsequent outcomes like hospitalisations and deaths. We further assume that cases only seek medical help once. |
| Sick people need hospitalisation | 1% | This estimate is highly uncertain. We have multiplied by 5 the percentage which has been observed for seasonal influenza in Germany (0.2%), to account for the apparent increased severity of COVID-19. The high uncertainty for this parameter is due to the likely under-diagnosis of mild cases in many settings (impacting the size of the denominator). It also may vary between countries given the use of hospital facilities to isolate mild cases. Modellers in the United Kingdom (UK) have used 4.4% (of all infected cases) [3], and for modelling in the United States 3%, 5% and 12% have been proposed [16]. The length of hospitalisation was assumed to be 10 days which is similar to other modelling work e.g., 10.4 days for the UK [3]. |
| Hospitalised cases need intensive care (ICU admission) | 25% | We used the data from a very large Chinese study for the ratio of “critical” to “severe” cases (i.e., $4.7\% / (13.8\% + 4.7\%) = 25.4\%$) [12]. This is similar to the Chinese case series reported by Wang et al. at 26.1% [17]. Nevertheless, it is higher than reported in a smaller case series from Singapore at 11% (2/18) [18]. A UK modelling study used a proportion of 30% “based on early reports from COVID-19 cases in the UK” [3]. |
| Intensive care cases requiring mechanical ventilation | 50% | We use the same value as per a US model of 50% [19] for additional calculations outside of CovidSIM. This proportion is around that reported in a Chinese study of 47% (17/36 ICU admissions) [17] but is less than in another Chinese study at 71% (37/52) [20]. |
| Sick people die from the disease (case fatality risk) | 0.45% | Given the relatively high quality of the healthcare systems in Europe, we considered the lower end of the range reported by the WHO for the infection fatality risk (IFR) of 0.3% to 1% (based on 3 publications) [21]. This IFR was then adjusted by the proportion assumed to be symptomatic (at 67%, as above) to give a case fatality risk (CFR) of 0.45% (i.e., $0.3\% \times 100\% / 67\% = 0.45\%$). Nevertheless, we note that higher estimates exist, including a CFR for “China outside of Hubei Province” of 0.81% (95%CI: 0.67 to 0.98; and adjusted for the time delay in reporting deaths) [22]. Another CFR for “China outside of Hubei Province” was similar, at 0.9% (95% credible interval: 0.6-1.3%; also adjusted for the time delay in reporting deaths) [23]. A value |
|  |  | used in UK modelling was an IFR of 0.9% [3]. |
| Basic reproduction number ($R_0$) | 1.5, 2.5 and 3.5 (3 scenario analyses) | On 6 March 2020, the WHO reported that this number was likely to be in the range of 2.0 to 2.5 [24]. But given persisting uncertainty, we used the same three values as in the modelling work by Hellewell et al [2]. Of note is that an earlier review of 12 studies [25], suggested estimates that ranged from 1.4 to 6.49, with a mean of 3.28, a median of 2.79 and interquartile range of 1.16. But this review also noted that in more recent studies, $R_0$ estimates seem to have stabilised at around 2–3. Recent UK modelling used an estimate of 2.4 (range: 2.0 to 2.6) [3]. |
| Relative contagiousness in the prodromal period | 50% | There is uncertainty around this value but we used the same estimate as in recent UK modelling [3]. This has biological plausibility as while there is similarity in viral loads between asymptomatic and symptomatic COVID-19 patients [26], it would be expected that those who are fully symptomatic (with a cough etc.) would be more likely to transmit infection. Of note is an estimate from the Diamond Princess cruise ship outbreak, that 17.9% of COVID-19 infections were from asymptomatic individuals (95% credible interval 15.5-20.2%) [27]. But it is unclear how generalisable this finding is given the crowded cruise ship conditions and the typically elderly nature of the passengers. |
| Latency period | 4 days | We used an average duration of 4 days as per Read et al [28], with a standard deviation of 25% (calculated using 16 stages; Erlang distribution). This is similar to the estimate in a Chinese study which reported a median latent period of 3.69 days [29]. |
| Prodromal period | 1 day | There is as yet insufficient data on this for COVID-19, so we used an assumed value for influenza (SD = 25%, Erlang distribution). |
| Symptomatic period | 10 days | The WHO-China Joint Mission report stated that “the median time from onset to clinical recovery for mild cases is approximately 2 weeks and is 3-6 weeks for patients with severe or critical disease” [8]. But given that mild cases may have been missed in this particular assessment, we used a slightly shorter time period of 10 days (SD = 25%, Erlang distribution). During this symptomatic period, cases were considered infectious. We note that there is evidence from COVID-19 cases of shedding of viral RNA from sputum that has outlasted the end of symptoms [30]. However, the significance of this for disease transmission is unknown. |
| **Interventions** |  |  |
| General contact reduction | Two scenarios (25%, 50%) and threshold analyses | This variable covers the summated impact of a potentially wide variety of different interventions: people may adopt enhanced personal hygiene measures (hand washing, cough etiquette etc.); they may decide to have fewer contacts (physical distancing); and governments may close venues and schools, restrict mass transit, curtail mass gatherings, and restrict travel (within and between countries).<br>**Scenario “25%”:** This scenario is our approximation of a modest level of the above listed interventions.<br>**Scenario “50%”:** This scenario assumed an intensification of the measures being adopted (relative to the above scenario).<br>**Threshold analyses:** This was where we increased the level of “general contact reduction” to a level which pushes the epidemic peak into the following year (i.e., past day 365 after the first introduction of the infection into the country). |
| Contact reduction begins | 19 February 2020 | This date is somewhat arbitrary but does represent a time of increased awareness of the disease in Europe as there were 9 European countries having reported cases to WHO and one reporting a death (in France). It is equivalent to day 5 of the simulated year. |
| Contact reduction duration | 6 months (9 months and “rest of year” in scenario analyses) | This period was arbitrary and was varied in threshold analyses (Table 1). As further discussed in the main text the feasibility of such sustained interventions for any country is highly uncertain and may not be realistic given the adverse social and economic implications. |
| Seasonality effect | Variation in $R_0$ of 25% | Winter conditions are known to accelerate transmission of influenza and also the other coronaviruses which cause common cold like symptoms [31]. Enveloped viruses show strong seasonality with winter peaks [32], and SARS-Cov-2 is an enveloped virus. Even though there are many uncertainties relating to seasonality and this novel coronavirus [33], it seems prudent to assume some seasonal fluctuation so we increased the average by 25% in winter and reduced it by 25% in summer (with a sinusoidal variation throughout the simulated year), using a mid-winter peak for Europe of 15 January (i.e., day 334 of the simulation). |
| Case isolation (only used in the threshold analyses) | Varied in threshold analyses (Table 1) | We set the following values in threshold analyses (while setting 0% for “general contact reduction” – see above):<br>• Probability that a sick person is isolated = varied in threshold analyses<br>• Maximum capacity of isolation wards = 4 per 10,000 population (i.e., see below).<br>• Contact reduction for cases in home isolation = 50% (this occurs while hospital isolation capacity is exceeded)<br>• Beginning of case isolation measures = the 19 February date as above (i.e., assuming increased clinician awareness from cases in nine European countries at this time).<br>• Duration of case isolation measures = 6 months (183 days), or 9 months (274 days) or the rest of the simulated year.<br>For isolation capacity in hospitals we assumed that 10% of hospital beds could be converted for this use during the pandemic, with the country having a total of 44,000 hospital beds; or 44 per 10,000 population. For this we used the median hospital beds per 1,000 population for 12 European states in the OECD and in the 5 to 15 million range for total population size, i.e., using data from: Austria, Belgium, Czechia, Denmark, Greece, Hungary, Norway, Portugal, Slovakia, Sweden and Switzerland [34]. If 10% of these hospital beds were used for isolation purposes, then this is 4 beds per 10,000 population for use in the modelling process, or 4,000 in total in the country. |

## References

1. World Health Organization. Coronavirus disease 2019 (COVID-19) Situation Report – 59. 2020;(19 March). https://www.who.int/docs/default-source/coronaviruse/situation-reports/20200319-sitrep-59-covid-19.pdf?sfvrsn=c3dcdef9_2.

2. Hellewell J, Abbott S, Gimma A, Bosse NI, Jarvis CI, Russell TW, et al. Feasibility of controlling COVID-19 outbreaks by isolation of cases and contacts. Lancet Glob Health. 2020.

3. Ferguson N, Laydon D, Nedjati-Gilani G, Imai N, Ainslie K, Baguelin M, et al. Impact of non-pharmaceutical interventions (NPIs) to reduce COVID-19 mortality and healthcare demand. Imperial College 2020;(16 March):1–20.

4. Wang C, Liu L, Hao X, Guo H, Wang Q, Huang J, et al. Evolving epidemiology and impact of non-pharmaceutical interventions on the outbreak of coronavirus disease 2019 in Wuhan, China. MedRxiv 2020;(6 March). https://www.medrxiv.org/content/10.1101/2020.03.03.20030593v1.

5. Lai S, Ruktanonchai N, Zhou L, Prosper O, Luo W, Floyd J, et al. Effect of non-pharmaceutical interventions for containing the COVID-19 outbreak: an observational and modelling study. MedRxiv 2020;(9 March). https://www.medrxiv.org/content/10.1101/2020.03.03.20029843v2.full.pdf.

6. World Health Organization. Novel Coronavirus (2019-nCoV) Situation Report – 5. 2020;(25 January). https://www.who.int/docs/default-source/coronaviruse/situation-reports/20200125-sitrep-5-2019-ncov.pdf?sfvrsn=429b143d_8.

7. Cohen J, Kupferschmidt K. Mass testing, school closings, lockdowns: Countries pick tactics in ‘war’ against coronavirus. Science 2020;(18 March). https://www.sciencemag.org/news/2020/03/mass-testing-school-closings-lockdowns-countries-pick-tactics-war-against-coronavirus.

8. WHO-China Joint Mission. Report of the WHO-China Joint Mission on Coronavirus Disease 2019 (COVID-19). 2020;(16-24 February). https://www.who.int/docs/default-source/coronaviruse/who-china-joint-mission-on-covid-19-final-report.pdf.

9. Kupferschmidt K, Cohen J. China’s aggressive measures have slowed the coronavirus. They may not work in other countries. Science 2020;(2 March). https://www.sciencemag.org/news/2020/03/china-s-aggressive-measures-have-slowed-coronavirus-they-may-not-work-other-countries.

10. Cowling B, Lim W. They’ve contained the coronavirus. Here’s how. New York Times 2020;(13 March). https://www.nytimes.com/2020/03/13/opinion/coronavirus-best-response.html.

11. Ansart S, Pelat C, Boelle PY, Carrat F, Flahault A, Valleron AJ. Mortality burden of the 1918-1919 influenza pandemic in Europe. Influenza Other Respir Viruses. 2009;3(3):99–106.

12. The Novel Coronavirus Pneumonia Emergency Response Epidemiology Team. The epidemiological characteristics of an outbreak of 2019 novel coronavirus diseases (COVID-19) — China, 2020. China CDC Weekly 2020. http://weekly.chinacdc.cn/en/article/id/e53946e2-c6c4-41e9-9a9b-fea8db1a8f51.

13. Reynolds DL, Garay JR, Deamond SL, Moran MK, Gold W, Styra R. Understanding, compliance and psychological impact of the SARS quarantine experience. Epidemiol Infect. 2008;136(7):997–1007.

14. Wu P, Fang Y, Guan Z, Fan B, Kong J, Yao Z, et al. The psychological impact of the SARS epidemic on hospital employees in China: exposure, risk perception, and altruistic acceptance of risk. Can J Psychiatry. 2009;54(5):302–311.

15. Wu J, Leung K, Bushman M, Kishore N, Niehus R, de Salazar P, et al. Estimating clinical severity of COVID-19 from the transmission dynamics in Wuhan, China. Nature Med. 2020;(E-publication 19 March).

16. Fink S. Worst-case estimates for U.S. coronavirus deaths. New York Times 2020;(Updated 14 March). https://www.nytimes.com/2020/03/13/us/coronavirus-deaths-estimate.html.

17. Wang D, Hu B, Hu C, Zhu F, Liu X, Zhang J, et al. Clinical characteristics of 138 hospitalized patients with 2019 novel coronavirus-infected pneumonia in Wuhan, China. JAMA. 2020;(E-publication 8 February).

18. Young BE, Ong SWX, Kalimuddin S, Low JG, Tan SY, Loh J, et al. Epidemiologic Features and Clinical Course of Patients Infected With SARS-CoV-2 in Singapore. JAMA. 2020.

19. Predictive Healthcare team at Penn Medicine. COVID-19 Hospital Impact Model for Epidemics. University of Pennsylvania, 2020. http://penn-chime.phl.io/.

20. Yang X, Yu Y, Xu J, Shu H, Xia J, Liu H, et al. Clinical course and outcomes of critically ill patients with SARS-CoV-2 pneumonia in Wuhan, China: a single-centered, retrospective, observational study. Lancet Respir Med. 2020;Published Online (21 February). https://doi.org/10.1016/S2213-2600(20)30079-5.

21. World Health Organization. Coronavirus disease 2019 (COVID-19) Situation Report – 30. 2020;(19 February). https://www.who.int/docs/default-source/coronaviruse/situation-reports/20200219-sitrep-30-covid-19.pdf?sfvrsn=3346b04f_2.

22. Wilson N, Kvalsvig A, Telfar Barnard L, Baker M. Case-fatality estimates for COVID-19 calculated by using a lag time for fatality. Emerg Infect Dis. 2020 [Early release 13 March]. https://doi.org/10.3201/eid2606.200320.

23. Mizumoto K, Chowell G. Estimating the risk of 2019 novel coronavirus death during the course of the outbreak in China, 2020. MedRxiv 2020;(23 February). https://www.medrxiv.org/content/10.1101/2020.02.19.20025163v1.

24. World Health Organization. Coronavirus disease 2019 (COVID-19) Situation Report – 46. 2020;(6 March). https://www.who.int/docs/default-source/coronaviruse/situation-reports/20200306-sitrep-46-covid-19.pdf?sfvrsn=96b04adf_4.

25. Liu Y, Gayle AA, Wilder-Smith A, Rocklov J. The reproductive number of COVID-19 is higher compared to SARS coronavirus. J Travel Med. 2020.

26. Zou L, Ruan F, Huang M, Liang L, Huang H, Hong Z, et al. SARS-CoV-2 Viral Load in Upper Respiratory Specimens of Infected Patients. New Engl J Med. 2020.

27. Mizumoto K, Kagaya K, Zarebski A, Chowell G. Estimating the asymptomatic proportion of coronavirus disease 2019 (COVID-19) cases on board the Diamond Princess cruise ship, Yokohama, Japan, 2020. Euro Surveill. 2020;25:pii=2000180. https://doi.org/2000110.2002807/2001560-2007917.

28. Read J, Bridgen J, Cummings D, Ho A, Jewell C. Novel coronavirus 2019-nCoV: early estimation of epidemiological parameters and epidemic predictions. MedRxiv 2020. doi: https://doi.org/10.1101/2020.01.23.20018549.

29. Li R, Pei S, Chen B, Song Y, Zhang T, Yang W, et al. Substantial undocumented infection facilitates the rapid dissemination of novel coronavirus (SARS-CoV2). Science (New York, NY. 2020.

30. Woelfel R, Corman V, Guggemos W, Seilmaier M, Zange S, Mueller M, et al. Clinical presentation and virological assessment of hospitalized cases of coronavirus disease 2019 in a travel-associated transmission cluster. MedRxiv 2020;(8 March). https://www.medrxiv.org/content/10.1101/2020.03.05.20030502v1.

31. Killerby ME, Biggs HM, Haynes A, Dahl RM, Mustaquim D, Gerber SI, et al. Human coronavirus circulation in the United States 2014-2017. J Clin Virol. 2018;101:52–56.

32. Price RHM, Graham C, Ramalingam S. Association between viral seasonality and meteorological factors. Sci Rep. 2019;9:929.

33. Cohen J. Why do dozens of diseases wax and wane with the seasons—and will COVID-19? Science 2020;(13 March). https://www.sciencemag.org/news/2020/03/why-do-dozens-diseases-wax-and-wane-seasons-and-will-covid-19.

34. OECD. Stat. Health care resources (2017 and 2018 data for hospital beds per 1000 population). https://stats.oecd.org/index.aspx?DataSetCode=HEALTH_REAC#.

